# Socio-demographic predictors of adherence to coronavirus disease prescribed recommendations and lockdown psychological impacts: Perspectives of Nigerian social media users

**DOI:** 10.1101/2020.09.09.20188482

**Authors:** Obasanjo Afolabi Bolarinwa, Olalekan Olagunju, Tesleem Babalola, Balsam Qubais Saeed

**Affiliations:** Discipline of Public Health Medicine, School of Nursing and Public Health, College of Health Sciences, University of KwaZulu-Natal, 4041, South Africa; Department of Demography & Social Statistics, Faculty of Social Sciences, Obafemi Awolowo University, Nigeria; Department of Clinical Sciences, College of Medicine, University of Sharjah, P.O. Box 27272, UAE; Sharjah Institute for Medical Research, University of Sharjah, P.O. Box 27272, Sharjah, UAE

**Keywords:** Coronavirus lockdown, Adherence to prescribed recommendations, Psychological impacts, COVID-19, Nigeria

## Abstract

**Background:** The coronavirus disease (COVID-19) pandemic is a highly infectious viral disease that has spread to over one hundred and eight countries, including Nigeria. Governments across the globe have been implementing preventive measures towards curbing the spread of the virus. These measures have continued to interfere with the general lifestyle of the people. Hence, this study was aimed at examining the socio-demographic predictors of adherence to prescribed recommendations and the psychological impacts of COVID-19 pandemic lockdown among Nigerian social media users.

**Methods:** This research implemented a cross-sectional survey using an online Google-based questionnaire to elicit required information from potential respondents via social media platforms. An external link to the questionnaire was shared among Nigerian social media users between for a month, and a total of 1,131 respondents participated in the survey. The explanatory and outcome variables were displayed by frequency and percentage distribution, while chi-square analysis was used to show the relationship between the explanatory and outcome variables at a 5% level of significance.

**Results:** The study showed that 99% of the respondents reported following some of the prescribed recommendations; however, only 40.4% of the respondents followed all the recommendations. More than three fifths (63.4%) of the respondents also reported that they feel stressed during the lockdown. All the selected socio-demographic characteristics were not predictors of the outcome variables as *p>0*.*05* except the professional background of the respondents *(P<0*.*05)*.

**Conclusion:** We concluded that most Nigerian social media users were complaining to the prescribed recommendations and that the younger age group, female respondents and respondents who are more educated had a higher proportion of reporting psychological impacts of lockdown.

## Introduction

### Background of the study

The disease known as a coronavirus (COVID-19) caused by SARS-CoV-2, has been designated a pandemic. This viral infectious disease is an acute respiratory illness (1), which has now spread across all the countries of the world (2, 3). Governments and medical officials are trying their best to curb the spread as much as possible (4). The virus is transmitted by aerosols that could remain suspended in the air for many minutes after coughing or sneezing or via close personal contact, such as touching or shaking hands with an infected person. These viruses may also spread when people touch contaminated objects or surfaces and their mouth, nose, or eyes. Moreover, it can remain viable for a few days on multiple surfaces (5, 6). All populations have been identified to be at the risk of COVID-19 irrespective of their socio-demographic characteristics or compositions; however, people with comorbidities and males older than 60 years have been reported to be more at risk of the virus (7).

The virus was declared by the World Health Organization (WHO) as a public health emergency of international concern on 31st January 2020, and on 11th March of the same year, WHO declared the outbreak a pandemic (4, 8). As of 2nd May 2020, over 3.3 million cases and 330,000 deaths have been reported globally (9), and these cases are still growing.

Gilbert and colleagues’ modelling study of the risk of COVID-19 importation from China indicates that the ability of African countries to manage the local transmission of the virus after importation hinges on implementing stringent measures of detection, prevention, and control (10). The country with the second-highest import risk ranking was Nigeria, with moderate capacity but high vulnerability and potentially significantly larger populations exposed to ineffective healthcare system (3).

The first reported case of the novel virus was imported into Nigeria in February 2020 by an Italian citizen (10), and the current number as at the time of this study showed that 2170 cases had been reported with 68 deaths in Nigeria (9). Since the vaccine is currently not available, it seems that the virus can only be slowed by extreme behavioral change and societal coordination, else the virus will spread within the country very fast (2). Preventative measures implemented by national, state, and local governments worldwide now affect the daily routines of millions of people worldwide, and these measures include social distancing and non-movement between and within countries (11-13). These measures are effective in slowing coronavirus spread and protect the health systems of the country (14).

Additionally, existing researches have demonstrated that the current most effective and efficient public health interventions are only feasible when the public duly accepts and adhere to the prescribed recommendations by the authorities (15). However, preliminary reports show vast differences in peoples’ willingness to practice measures that can reduce the virus transmission (2, 16). For instance, a study conducted in Italy concluded that the majority of the respondents adherend to nationally prescribed health measures towards curbing the spread of the virus (17). In the same vein, a sentiment study conducted on compliance to lockdown in India showed a positive curve towards the compliance despite the negativity, fear, disgust, and sadness about the lockdown (4). It’s expected that due to the high cost of complete isolation and healthcare services, compliance with prescribed recommendations and national lockdown to strategically reduce contact is expected to be higher but this is without consequences or implications to mental health well-being of the people (15).

During the lockdown, people were asked to stay at home by socially and physically isolate themselves to prevent being infected (18). Although these measures are effective in preventing the uncontrollable spreading of the virus, on the other hand, these measures can negatively affect the mental health of the population (19), and relaxation will almost certainly trigger a further epidemic wave of deaths (20). Studies have shown that social separation or quarantine of non-infected persons for an extended period may have adverse effects, such as loneliness, a rise in fear and anxiety which may have implications on mental health (12-16, 21). The previous outbreaks of another family of coronaviruses such as middle east respiratory syndrome coronavirus (MERS-CoV) and severe acute respiratory syndrome coronavirus 2 (SARS-Cov-2) had been linked to anxiety, depression, and psychological challenges(22); also, that the fear of the unknown raises anxiety levels in healthy individuals and those with pre-existing mental health conditions are more at risk (23).

There has been a global rise recently in the spread of misinformation that has plagued the scientific community and the public (24). In our current digital world, online platforms are perhaps the most accessible source of health-related information for the public (25). As more social interactions move online, the conversation around COVID-19 has continued to expand, with growing numbers turning to social media for both information sourcing and social interaction globally and in Nigeria (11). Twitter and other social media platforms are essential sources of breaking news around the globe (26). However, limited studies have been able to link the socio-demographic predictors of compliance to prescribed recommendations and COVID-19 lockdown psychological impacts among social media users. Thus, this study examined the socio-demographic predictors of adherence to prescribed recommendations and psychological impacts of COVID-19 among Nigerian social media users.

## Design and Methods

### Participant settings

The current estimated population of Nigerian social network users stood at 28.15 million, which accounts for about 14 percent of the total population, and it is projected to reach 44.62 million by the year 2025(27). Social media users were selected as the study population due to the current restrictions on movement and physical interaction in the country. This cross-sectional survey used a Google-based, anonymous online questionnaire to gather data from respondents via social media platforms such as Telegram, Instagram, Facebook, WhatsApp, and Twitter. On these platforms, the google based questionnaire link was shared among the participants. A snowball sampling technique was adopted to involve more social media users currently residing in Nigeria during the COVID-19 pandemic lockdown by telling those who first got the external google based questionnaire link to share with their contacts. This unidentified online survey was conducted for one month (between 1 April to 31st May 2020), and a total of 1,131 participants were involved.

### Procedure

Due to the social distancing rules imposed by the Nigerian Government and the enforcement of curfew/lockdown, physical interaction was not feasible, so the study survey was promoted online via social media platforms, and existing study participants were encouraged to share the online google based questionnaire with potential respondents. Participation was completely consensual, voluntary and anonymous. All respondents were asked an informed consent question at the beginning of the questionnaire by asking if they were interested in participating in the online google based questionnaire for this study or not, those who chose that they are not interested in participating were signed-out from completing the next phase of the online google based questionnaire while those who agreed to participate were allowed to move to the next phase involving the completion of the online google based questionnaire. The online google based questionnaire elicited socio-demographic variables such as gender, age, educational attainment and professional background of the respondents, while outcome variables such as compliance to prescribed recommendations and psychological impacts of lockdown during the lockdown among Nigerian social media users.

### Variables Definition

Respondent age, gender or sex, educational level, and professional background variables were explanatory variables. The outcome variables were recommendation compliance, feeling regarding the COVID-19 pandemic and respondents’ adaptation. Recommendation compliance refers to whether respondents comply with the country’s ministry of health recommendations, while feelings regarding the COVID-19 pandemic refers to the respondent’s feelings during the lockdown. Respondents were asked about their opinions regarding the COVID-19 epidemic; those that reported nervous/anxious, fear, angry, lonely, and bored were coded stressed, while those that said just fine, happy, and relaxed/optimistic were coded not stressed. Respondents’ adaptations refer to coping strategies used by the respondent. Respondents were coded adapting well, if engaged in positive activities like watching television, reading books/magazines, volunteering, working from home, etc. Respondent was coded not adapting well if they engaged in harmful activities like fighting with everyone, talking to themselves, having problem sleeping, etc.

### Data Analysis

The data collected were analysed using Stata 14 statistical tool. Findings were described in table and figure formats, using frequencies and percentages to explain specific variables of the sample population. A bivariate analysis (chi-square test) was conducted to predict the influence of socio-demographic factors on the outcome variables. Results with a p-value of less than 0.05 were significant predictors in the bi-variate analysis.

### Ethical Consideration

The ethics committee of Obafemi Awolowo University, Nigeria, approved the study of Obafemi Awolowo University Nigeria. Participants’ permission was sought before filling out the online google based questionnaire. All those who agreed to participate in the survey were granted access to the online form.

## Results

Table 1 shows the socio-demographic distribution of the respondents. The table revealed that more than 77.6 percent of respondents were between the ages of 18 to 39, while the remainder were between the ages of 40 and above. Sex reveals 42.1% were male, and 57.9% were female. The table shows that respondents were well educated, with 5.7% without a college education. More than half of the respondents were from a scientific/medical professional background (61.3%).

**Table 1:** Respondents’ socio-demographic characteristics.

| Variable | N=1131 | % |
| --- | --- | --- |
| **Sex** |  |  |
| Male | 474 | 42.1 |
| Female | 652 | 57.9 |
| **Educational level** |  |  |
| No college (formal education) | 64 | 5.7 |
| College education | 1067 | 94.3 |
| **Age range** |  |  |
| Below 40 years | 878 | 77.6 |
| 40+ | 253 | 22.4 |
| **Professional Background** |  |  |
| Non-scientific/non-medical | 438 | 38.7 |
| Scientific/medical | 693 | 61.3 |

Table 2 presents the respondents’ compliance with prescribed recommendations. The table shows that 9 out of every respondent reported that they follow the country ministry of health recommendations. When asked about the extent they follow the recommendations, 4 out of every ten reported that they follow all the recommendations. Respondents were asked how frequently they touch their face; only 3.8% reported that they never touch their face, 34.3% rarely touch their face, 34.7% touches their face sometimes, 21.2% touches their face often, and only 6% touch their face always.

**Table 2:** Prescribed recommendation compliance.

| Variable | N=1131 | % |
| --- | --- | --- |
| **Do you follow the recommendation of your health ministry/government** |  |  |
| No | 11 | 1.0 |
| Yes | 1120 | 99.0 |
| **To which extent do you follow them** |  |  |
| Not at all | 4 | 0.3 |
| I follow some but not all | 150 | 13.3 |
| I follow most of them | 520 | 46.0 |
| I follow all the recommendations | 457 | 40.4 |
| **How frequently do you touch your face** |  |  |
| Never | 43 | 3.8 |
| Rarely | 388 | 34.3 |
| Sometimes | 392 | 34.7 |
| Often | 240 | 21.2 |
| Always | 68 | 6.0 |

Table 3 presents the respondents’ psychological impacts and coping strategies & source of information. Six out of every ten respondents reported that they were stressed, and about four reported that they were not stressed. Concerning coping and adaptation during the lockdown, 83.7% reported that they are adapting well by watching TV/movies, spending time with family, reading books/magazines, and working from home. Source of information about the COVID-19 pandemic revealed that 8 out of every ten respondents heard the information from social media (Facebook, Instagram, WhatsApp profile, etc.) while the remaining heard from TV (14.1%), friends/family (2.4%), newspaper (2.0%) and other sources (1.4%).

**Table 3:**
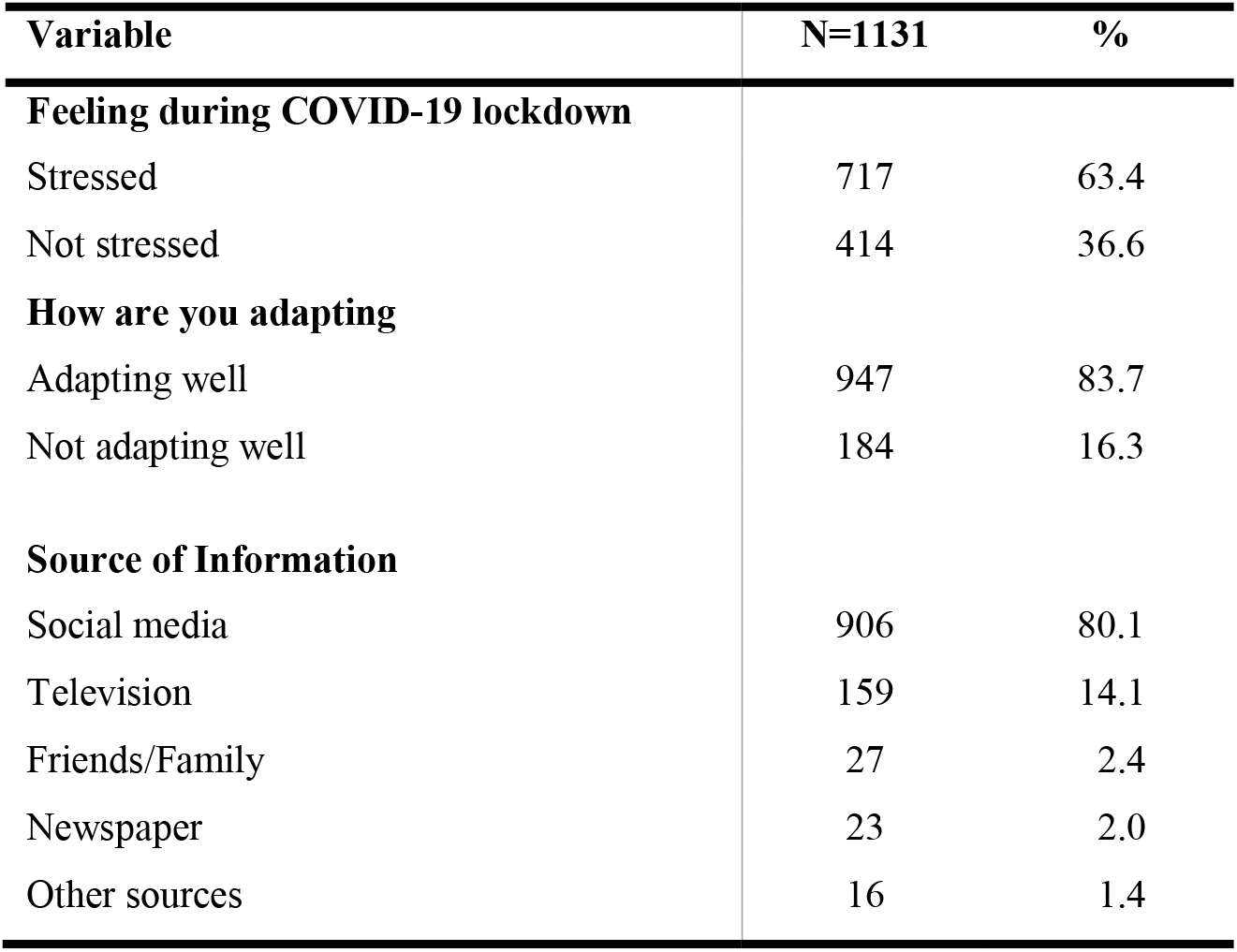
Lockdown psychological impacts & Source of Information.

Table 4 below presents the recommendations compliance and respondent socio-demographics. The table reveals that as age increases, the number of respondents’ compliance with recommendations decreases, but almost all the respondents in both categories adhere to recommendations. There is no statistically significant relationship between the age of respondents, gender, level of education, and respondents’ professional background and compliance with the recommendations.

**Table 4:**
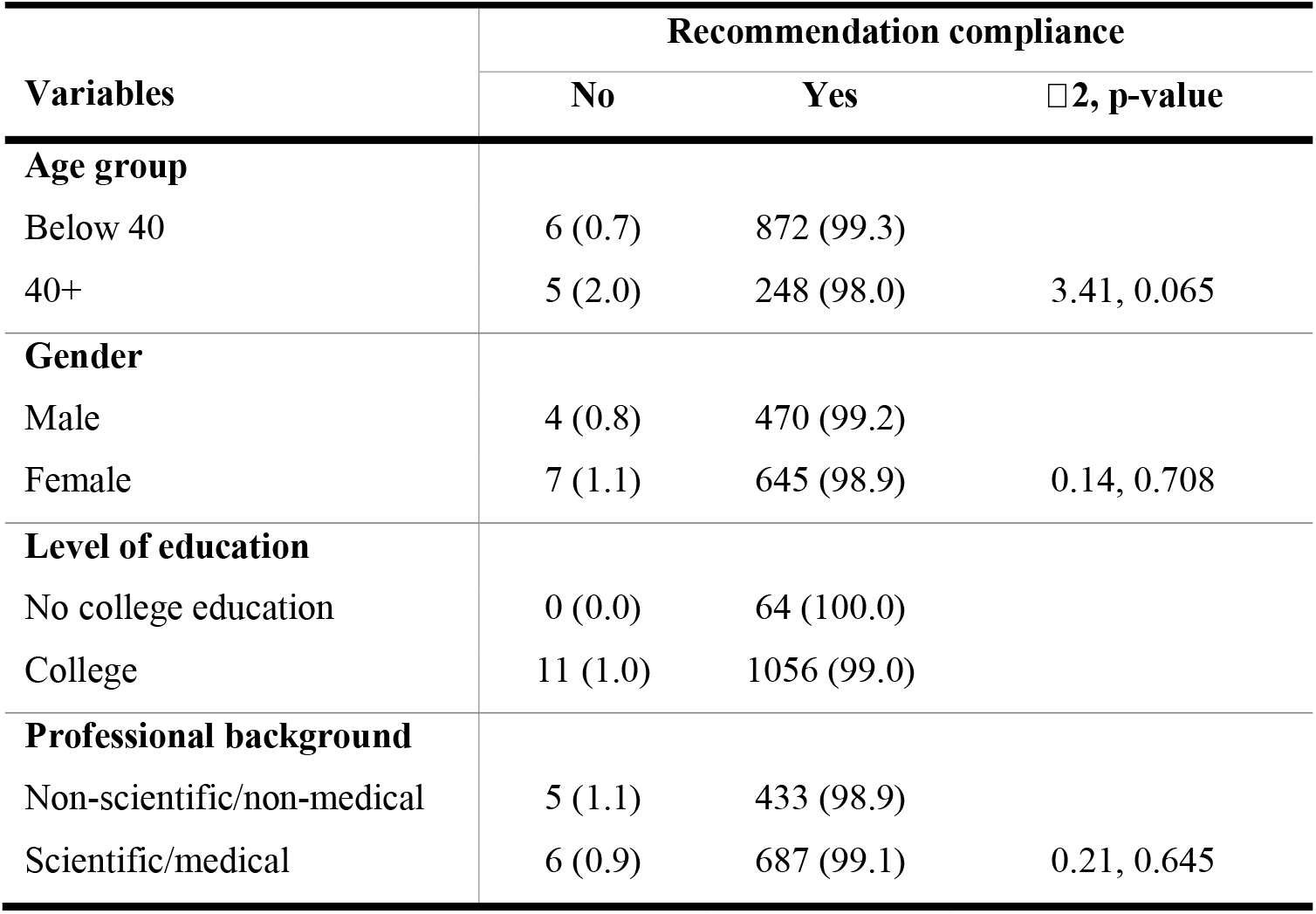
Association between socio-demographic and recommendation compliance.

Table 5 describes the association between respondents’ socio-demographic variables and feelings during the COVID-19 lockdown. More than half of respondents in both age categories reported been stressed during the lockdown. The same trend was observed for the level of education. More than half of respondents with no college education and college education feel stressed during the pandemic lockdown, but this is not statistically significant in this study. The gender and professional background of the respondent also showed no statistically significant.

**Table 5:** Association between socio-demographic and Feeling during COVID-19 lockdown.

| Variables | Feeling during COVID-19 lockdown |  |  |
| --- | --- | --- | --- |
| | Stressed | Not stressed | $\chi^2$, p-value |
| **Age group** |  |  |  |
| Below 40 | 565 (64.4) | 313 (35.4) |  |
| 40+ | 152 (60.1) | 101 (39.9) | 1.54, 0.214 |
| **Gender** |  |  |  |
| Male | 307 (64.8) | 167 (35.2) |  |
| Female | 410 (62.7) | 247 (37.3) | 0.66, 0.416 |
| **Level of education** |  |  |  |
| No college education | 36 (56.3) | 28 (43.7) |  |
| College education | 681 (63.8) | 386 (36.2) | 150, 0.222 |
| **Professional background** |  |  |  |
| Non-scientific/non-medical | 290 (66.2) | 148 (33.8) |  |
| Scientific/medical | 427 (61.6) | 266 (38.4) | 2.44, 0.118 |

Table 6 presents the respondent’s socio-demographic and adaptation during the pandemic. Across all socio-demographics, it was observed that the majority of respondents were adapting well. The table shows that about 8 out of every ten respondents in both age group categories were adapting well. The age group of respondents, gender and level of education were not statistically significant. The table revealed that about 8 out of every ten respondents in the education categories reported that they were adapting well. The professional background of the respondent shows a statistically significant relationship in that more of the respondents with a scientific/medical background were adapting well.

**Table 6:** Association between socio-demographic and respondents’ adaptation.

| Variables | Respondents' adaptation |  |  |
| --- | --- | --- | --- |
|  | Adapting well | Not adapting well | χ<sup>2</sup>, p-value |
| **Age group** |  |  |  |
| Below 40 | 739 (84.2) | 139 (15.8) |  |
| 40+ | 208 (82.2) | 45 (17.8) | 0.55, 0.458 |
| **Gender** |  |  |  |
| Male | 405 (85.4) | 69 (14.6) |  |
| Female | 542 (82.5) | 115 (17.5) | 1.75, 0.185 |
| **Level of education** |  |  |  |
| No college education | 54 (84.4) | 10 (15.6) |  |
| College education | 893 (83.7) | 174 (16.3) | 0.02, 0.886 |
| **Professional background** |  |  |  |
| Non-scientific/non-medical | 346 (79.0) | 92 (21.0) |  |
| Scientific/medical | 601 (86.7) | 92 (13.3) | 11.77, 0.001** |

## Discussion

This study was carried out to examine the socio-demographic predictors of adherence to the prescribed recommendation and psychological impacts of lockdown among Nigerian social media users. As the world faces the coronavirus threat, many commentators and national leaders worldwide are beginning to recognize this as a genuine threat to national security(28). The danger of spreading COVID-19 does not come from an external adversary, but from peoples’ behaviour(29), adherence to prescribed recommendations has been studied to positively affect the spreading of the virus(4),(17). The same level of compliance was observed across all the selected socio-demographic characteristics in this study, as most of the respondents reported having been complying with the recommendations prescribed by the Nigerian Government. However, none of the selected socio-demographic characteristics of the respondents was a predictor of adherence to prescribed recommendations in this study.

It is crucial to know how people adapt to their various environments during COVID-19 lockdown in Nigeria; thus, this study further examined the socio-demographic predictors of the psychological impacts of COVID-19 lockdown on Nigerian social media users. The findings from this study shows that respondents who were younger and active were adjusting well. This may be because they have the energy to engage in different activities. Also, young people are the best with technology, and with the internet, they can relate with their friends in any part of the world. The educational level category also shows that respondents who were more educated were adapting well. This is because they are more knowledgeable and can easily understand the situation than those with no formal education. The respondents’ professional background was another socio-demographic characteristic we explore on adaptation during the lockdown in this study. The finding shows that respondents with scientific/medical professional backgrounds were more likely to adapt well, which may be because of their awareness of preventive measures.

The study also assesses the psychological impacts of lockdown on respondents’ socio-demographic characteristics on whether they are stressed or not. The finding shows that younger age groups, female respondents, educated respondents and respondents with professional scientific background reported having been stressed during coronavirus lockdown in Nigeria. It’s paradoxical because these same categories of socio-demographic characteristics reported having been adapting well based on the activities they were engaging with during lockdown; however, among the selected socio-demographic characteristics of the respondents, only professional background shows to be a predictor of psychological impacts of COVID-19 lockdown among Nigerian social media users. It’s pertinent to know that this same level of variation was reported previously in other studies with the same focus on other coronavirus family outbreaks such as MERS-CoV and SARS-CoV-2(22, 30).

### Strengths and limitations

The main strength of this study is the ability to capture data of interest during a pandemic without any physical contact with any of the respondents, and this is also linked to the fact that answering the questions was done at the convenience of the respondents without any interference. However, this study is also susceptible to certain limitations. The study data collection was done via social media platforms, as such, the younger population were more than the older population in this study, and this is because a larger number of the younger population are more acquitted to the use of technology than the older population hence, the study findings doesn’t depict the true picture of Nigeria populations as a whole.

## Conclusion

The study concluded that the majority of Nigerian social media users were adhering to the prescribed recommendations outlined by the government or health minister. As much as some selected socio-demographic characteristics of Nigerian social media users were not predictors of adherence to prescribed compliance, feelings and adaptation during COVID-19 lockdown, some sub-categories of the socio-demographic characteristics such as younger age group, female respondents, educated respondents and respondents with professional scientific backgrounds showed that a higher proportion of the respondents reported feeling stressed during COVID-19 lockdown in Nigeria. Psychological programmes that will consider the mental health of these sub-categories of respondents should be promoted in Nigeria.

The study further concluded that among the selected socio-demographic characteristics, only the professional background of the respondents was the major predictor of the psychological impact of COVID-19 lockdown among Nigerian social media users, as such an intervention that will address mental health and wellbeing of Nigerians during COVID-19 pandemic or future pandemic should be targeted at Nigerian social media users with a professional scientific background.

### Significance for public health

The ongoing novel coronavirus 2019 (COVID-19) pandemic has significantly changed the public’s day-to-day activities. To curb the virus’s spread, several health prescribed recommendations were imposed by the government, such as social distancing and national lockdown. Some populations with specific socio-demographic characteristics reported that the nation-wide lockdown had affected their behaviour and interaction with other people; thus, there is a need to examine the socio-demographic predictors of adherence to prescribed recommendations and psychological impacts of COVID-19 lockdown. The study findings showed that a higher significant proportion of the younger population, female population, educated population, and population with scientific professional background were psychologically affected by COVID-19 lockdown in Nigeria, which could have an adverse effect on their mental health well-being. Hence intervention to mitigate such adverse effects should be targeted at the population with the stated socio-demographic characteristics.

## Data Availability

Data is available from the authors after a reasonable request.

## Acknowledgements

We acknowledge all the volunteers who participated in the online survey to make this study possible.

